# MicroRNA-mRNA networks define translatable molecular outcome phenotypes in osteosarcoma

**DOI:** 10.1101/19007740

**Authors:** Christopher E. Lietz, Cassandra Garbutt, William T. Barry, Vikram Deshpande, Yen-Lin Chen, Santiago A. Lozano-Calderon, Yaoyu Wang, Brian Lawney, David Ebb, Gregory M. Cote, Zhenfeng Duan, Francis J. Hornicek, Edwin Choy, G. Petur Nielsen, Benjamin Haibe-Kains, John Quackenbush, Dimitrios Spentzos

**Affiliations:** Department of Orthopaedic Surgery, Massachusetts General Hospital, Harvard Medical School, Boston, MA; Department of Biostatistics and Computational Biology, Dana Farber Cancer Institute, Harvard Medical School, Boston, MA; Department of Pathology, Massachusetts General Hospital, Harvard Medical School, Boston, MA; Department of Radiation Oncology, Massachusetts General Hospital, Harvard Medical School, Boston, MA; Department of Biostatistics, Harvard School of Public Health, Boston, MA; Pediatric Hematology-Oncology, Massachusetts General Hospital, Harvard Medical School, Boston, MA; Department of Hematology/Oncology, Massachusetts General Hospital, Harvard Medical School, Boston, MA; Department of Orthopaedic Surgery, UCLA, Los Angeles, CA; Department of Medical Biophysics, Princess Margaret Cancer Centre, University of Toronto

**Keywords:** Osteosarcoma, microRNA, prognostic signature, sequencing, methylation, pharmacogenomics

## Abstract

**Background:** There is a lack of well validated biomarkers in osteosarcoma, a rare, recalcitrant disease with variable outcome and poorly understood biologic behavior, for which treatment standards have stalled for decades. The only standard prognostic factor in osteosarcoma remains the amount of pathologic necrosis following pre-operative chemotherapy, which does not adequately capture the biologic complexity of the tumor and has not resulted in optimized patient therapeutic stratification. New, robust biomarkers are needed to understand prognosis and better reflect the underlying biologic and molecular complexity of this disease.

**Methods:** We performed microRNA sequencing in 74 frozen osteosarcoma biopsy samples, the largest single center translationally analyzed cohort to date, and separately analyzed a multi-omic dataset from a large (n = 95) NCI supported national cooperative group cohort. Molecular patterns were tested for association with outcome and used to identify novel therapeutics for further study by integrative pharmacogenomic analysis.

**Results:** MicroRNA profiles were found predict Recurrence Free Survival (*5-microRNA profile*, Median RFS 59 vs 202 months, log rank p=0.06, HR 1.87, 95% CI 0.96-3.66). The profiles were independently prognostic of RFS when controlled for metastatic disease at diagnosis and pathologic necrosis following chemotherapy in multivariate Cox proportional hazards regression (*5-microRNA profile*, HR 3.31, 95% CI 1.31–8.36, p=0.01). Strong trends for survival discrimination were observed in the independent NCI dataset, and transcriptomic analysis revealed the downstream microRNA regulatory targets are also predictive of survival (median RFS 17 vs 105 months, log rank p=0.007). Additionally, DNA methylation patterns held prognostic significance. Through machine learning based integrative pharmacogenomic analysis, the microRNA biomarkers identify novel therapeutics for further study and stratified application in osteosarcoma.

**Conclusions:** Our results support the existence of molecularly defined phenotypes in osteosarcoma associated with distinct outcome independent of clinicopathologic features. We validated candidate microRNA profiles and their associated molecular networks for prognostic value in multiple independent datasets. These networks may define previously unrecognized osteosarcoma subtypes with distinct molecular context and clinical course potentially appropriate for future application of tailored treatment strategies in different patient subgroups.

## BACKGROUND

Osteosarcoma is a primary bone malignancy most prevalent in adolescents and young adults, with a second peak in later adulthood. While chemotherapy with the standard cisplatin/doxorubicin/methotrexate regimen (known as MAP), combined with surgical resection has markedly improved prognosis in patients with localized tumors, up to 40-50% of patients experience relapse and eventually succumb to the disease and the prognosis for patients presenting with metastatic disease is even poorer [1],[2]. There have been no new therapeutic advances in the last 30 years and progress in this disease is hindered by the lack of well validated biomarkers of outcome that may facilitate stratification of patients for new therapies [3],[4],[5],[6]. Pathologic necrosis in response to neoadjuvant chemotherapy holds prognostic significance but is imperfectly correlated with outcome in the subset of patients with suboptimal response [2]. In addition, it is semi-quantitative, and can only be assessed by expert pathologists only after several cycles of chemotherapy are already administered. A recent large randomized international trial (EURAMOS [6]), using pathologic necrosis to stratify patients for the addition of ifosfamide/etoposide or interferon to the adjuvant treatment, failed to produce a survival benefit underscoring both the need for novel therapeutics as well as the limitations of currently available prognostic/stratification markers leading to an increased interest in molecular markers of outcome. In this context, miRNAs have recently gained attention, due to their recognized regulatory role for large numbers of downstream genes in cancer. Our group previously presented pilot findings suggesting that microRNAs are useful in osteosarcoma prognostic stratification [7]. Here, we present an external, large-scale validation study and reproducibility assessment of previously defined miRNA profiles prognostic of outcome, using two independent large patient cohorts. One is a single center cohort from a large referral center (Massachusetts General Hospital), and the other is based on data recently released by the NCI Therapeutically Applicable Research to Generate Effective Treatments (TARGET) program, analyzed by RNA sequencing and Real Time PCR, respectively, and representing the largest translationally studied clinical cohorts to date, in this rare tumor. We further showed that gene targets of the prognostic miRNAs also provide robust outcome prediction and suggest that the miRNAs potentially define new, previously unrecognized molecular phenotypes with clinical relevance. Finally, we explore possible novel therapeutic implications by analyzing connections of the molecular profiles with new drugs that may be effective in osteosarcoma.

## RESULTS

### Characteristics of the two osteosarcoma cohorts and description of candidate miRNA signatures

The clinical and pathologic characteristics of the two osteosarcoma cohorts are shown in Table 1. In general, the NCI TARGET cohort included patients with relatively younger age and was more recently initiated thus providing shorter follow up than the MGH cohort. All but 10 MGH samples were pre-chemotherapy diagnostic biopsy samples, the remaining being pre-chemotherapy diagnostic resections. For the NCI TARGET dataset, all samples were pre-chemotherapy, but information on how many were biopsies or pre-chemotherapy diagnostic resections is not available. Virtually all patients in the MGH cohort had undergone treatment with at least the standard MAP (or AP) regimen. Full individual patient detailed treatment information is not yet available for the NCI TARGET cohort, but the reported treatment protocols contained at least the three components of the MAP regimen, (doxorubicin, cisplatin and methotrexate).

**Table 1.** Clinical characteristics of the MGH and TARGET cohorts.

| **Cohort** | **MGH** | **TARGET <sup>1</sup>** |
| --- | --- | --- |
| N | 74 | 95 <sup>2</sup> |
| Years collected | 1994-2013 | 2000-2012 |
| before 2000 | 25 | 0 |
| 2000-2004 | 23 | 25 |
| 2005-2009 | 19 | 45 |
| after 2009 | 7 | 22 |
| Not available | 0 | 3 |
| Age at Diagnoses (years) |  |  |
| Median | 22.5 | 14.7 |
| Range | 8 - 72 | 3-39 |
| Sex (# / %) |  |  |
| Male | 49 / 66 | 54 / 57 |
| Female | 25 / 34 | 41 / 43 |
| Tumor Location |  |  |
| Axial | 2 | 5 |
| Appendicular | 72 | 90 |
| Upper | 7 | 7 |
| Lower | 65 | 83 |
| Histologic Subtype |  |  |
| NOS | 42 | Not available |
| Osteoblastic | 5 |  |
| Fibroblastic | 4 |  |
| Chondroblastic | 16 |  |
| Small Cell | 4 |  |
| Mixed | 3 |  |
| Metastases at Diagnoses (# / %) |  |  |
| No | 57 / 77 | 71 / 75 |
| Yes | 17 / 23 | 24 / 24 |
| Chemotherapy regime |  |  |
| MAP or AP | 29 | 17 |
| MAP and other | 40 | 9 |
| Other | 3 | 0 |
| Not available | 2 | 69 |
| Chemo-response<sup>3</sup> |  |  |
| Optimal | 25 | 22 |
| Suboptimal | 26 | 30 |
| Not available | 23 | 43 |
| Events |  |  |
| Deaths <sup>4</sup> | 28 | 37 <sup>5</sup> |
| Recurrences | 36 | 47 <sup>6</sup> |
| Follow Up Time (years) |  |  |
| Median | 6.4 | 3.6 |
| Alive at last follow up | 44 | 58 |
| AWD | N = 11 | Not available |
| Avg. Follow Up and range | 10.5 (4.3 – 19.0) |  |
| NED | N = 33 | Not available |
| Median follow up and range | 11.2 (2.8 – 19.2) |  |

<sup>1</sup> Data collection is ongoing by the NCI for the TARGET cohort. <sup>2</sup>N<sub>miRNA</sub> = 86, N<sub>RNAseq</sub> = 93, N<sub>Methylation</sub> = 83. <sup>3</sup>Optimal chemo-response was defined as $\geq 90\%$ tumor necrosis at definitive surgery. <sup>4</sup>Deaths from osteosarcoma. <sup>5</sup>30 deaths in the TARGET miRNA cohort. <sup>6</sup>41 recurrences in the TARGET miRNA cohort.

We mapped our previously published prognostic signatures to the new cohorts for further validation. These signatures were a *5-miRNA* profile (hsa-miR-495-3p, hsa-miR-487b-3p, miR-410-3p, hsa-miR-329-3p, hsa-miR-664a-3p), and a *22-miRNA* profile (which includes the 5 miRNAs, Table S1) which had been previously defined in an initial pilot discovery sample cohort [7], that was entirely separate from any of the two cohorts described in this study and was studied using a different assay technology (expression microarray). It has been recognized that the majority of the previously identified prognostic miRNAs are encoded on the 14q32 chromosomal locus, which is the largest non-coding cluster in the human genome and is regulated partly via methylation and genomic imprinting mechanisms [8],[9],[10],[11]. Therefore, secondarily, we considered an expanded set of 27 miRNAs located on 14q32 that were previously found to be univariately prognostic of outcome in the pilot dataset [8]. Due to mapping attrition across platforms, from the 22-miRNA profiles, 21 and 18 transcripts were mapped onto the MGH RNAseq, and TARGET qRTPCR data, respectively. Similarly, from the 27-miRNA profile, 26 transcripts were mapped onto the TARGET Taqman qRTPCR data.

### Application of the prognostic miRNA profiles in the MGH osteosarcoma cohort

We tested the prognostic value of the signatures on the MGH small RNA sequencing dataset (n=74), after excluding samples with no or very low tumor content in the frozen specimen by pathology review. Given the technical differences and mapping attrition between the different assay platforms, an application of a fully pre-defined prognostic regression model could not be performed, thus we tested our previously identified signatures with two approaches that are less sensitive to the technical differences between the different assays: First, we performed unsupervised hierarchal clustering with the two miRNA profiles, which provided strong prognostic discrimination (Fig. 1a-b). Specifically, the 5-miRNA profile and 22-miRNA profiles discriminated between two groups with median RFS 59 vs 202 months, (log rank p=0.06, HR 1.87, 95% CI 0.96-3.66), and 33 months vs not reached, (log rank p=0.032, HR 1.99, 95% CI 1.04-3.80), respectively. Cluster reproducibility was high as indicated by the cluster R index [12] (5-miRNA profile R: 0.80, 22-miRNA profile R: 0.94). Further, we performed individualized patient prediction via supervised analysis using the “signed averaged expression” survival prediction method. This is a very simple approach to achieve an event risk score for each patient via averaging expression levels of candidate markers weighted by their Hazard Ratio for association with outcome, greatly minimizing the overfitting risk associated with more complex models, as we and others have previously shown [8]. With this approach, and using signs based on the pre-defined Hazard Ratios form the previously published “discovery” dataset [7] we also showed a robust prognostic performance of the previously defined signatures (Fig. 1c-d-e-f). While the analysis using the entire cohort did not produce significant prognostic discrimination, an analysis stratified by the presence of metastatic tumor at the time of diagnosis resulted in significantly different prognostic groups. Specifically, both profiles predicted two groups with median RFS 202 months vs not reached (for the group without metastasis at diagnosis) and median RFS 13 vs 27 months (for the group with metastasis at diagnosis) by Kaplan Meier analysis (stratified log rank p=0.031 and 0.042, for the 5-miRNA and 22-miRNA profiles, respectively).

**Figure 1.**
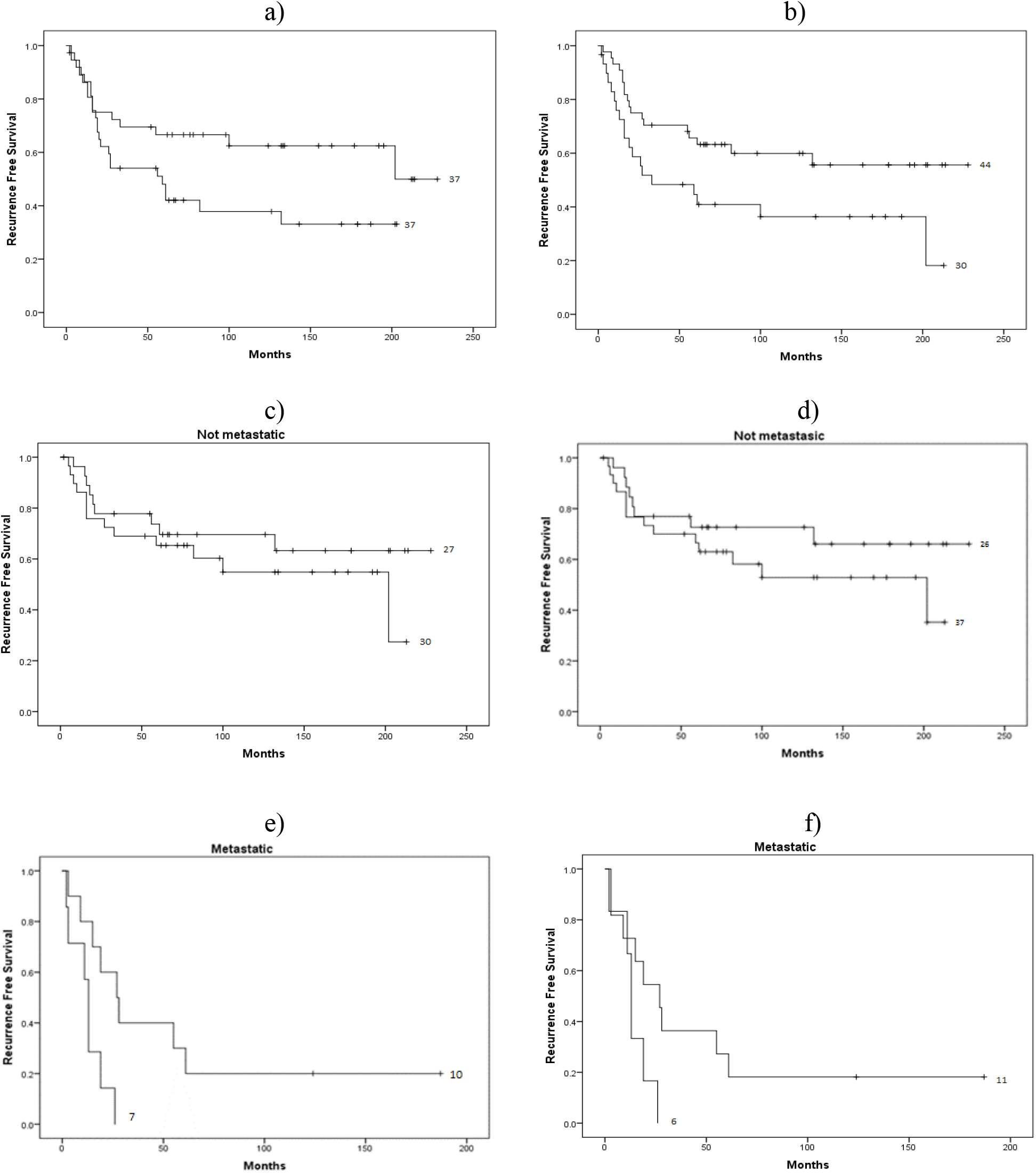
Kaplan-Meier log-rank Recurrence Free Survival analysis of candidate profiles on the MGH dataset. <u>Unsupervised hierarchal clustering</u> a) *5-miRNAs*, Median RFS 59 vs 202 months, p=0.06, b) *22 miRNAs*, 33 months vs not reached, p=0.032. <u>Supervised signed average prediction:</u> *5 miRNAs*, stratified p=0.031, median RFS: c) Not metastatic: 202 months vs not reached, e) Metastatic: 13 vs 27 months. *22 miRNAs*, stratified p=0.042, median RFS: d) Not metastatic: 202 months vs not reached, f) Metastatic: 13 vs 27 months.

The profiles were also prognostic of Overall Survival and discriminated between two clustering-based groups with median OS 69 months vs not reached, (log rank p=0.012, HR 2.65, 95% CI 1.20-5.89), and median OS 100 months vs not reached, (log-rank p=0.06, HR 2.00, 95% CI 0.95-4.22), for the 5-miRNA and 22-miRNA profiles, respectively (Fig. 2). In multivariate analysis, both profiles maintained independent prognostic value adjusted for metastatic status at diagnosis (HR 2.28, Cox regression p=0.04 and HR 2.00, p=0.06 for the 5-miRNA and the 22-miRNA profile, respectively). The 5-miRNA profile also maintained prognostic value even when controlling for both metastatic status and pathologic necrosis, despite the limited statistical power of such a multivariate analysis (HR 6.1, 95% CI 1.90-19.87). Analysis using the supervised signed average expression method produced similar results with a strong trend for survival (Fig. S1).

**Figure 2.**
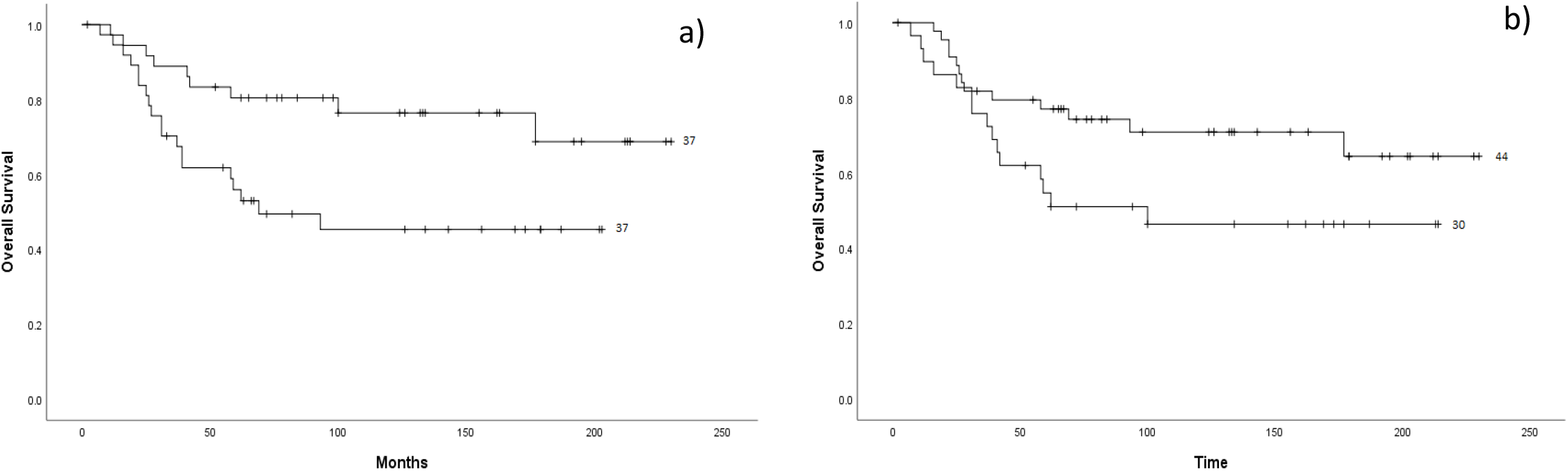
Kaplan-Meier log-rank Overall Survival analysis of candidate profiles on the MGH dataset by unsupervised hierarchical clustering. a) *5-miRNAs*, Median OS 69 months vs not reached, p=0.012, b) *22 miRNAs*, 100 months vs not reached, p=0.061.

The prognostic power of the profiles was found to be independent of confounding by known major clinicopathologic prognostic factors. For example, as above, the 5-miRNA profile was significantly prognostic of RFS when stratified by the presence of metastasis at diagnosis (no metastasis: median RFS 202 vs not reached, metastasis: 13 vs 27 months, p=0.031). In multivariate Cox proportional hazards regression, the miRNA signatures remained independently prognostic of RFS when controlled for the presence of metastatic disease at diagnosis and pathologic necrosis following preoperative chemotherapy (5 miRNA profile: HR 3.31, 95% CI 1.31-8.36, p=0.01).

Finally, we tested the secondary, (also predefined) 27 miRNA profile, consisting of the subset of miRNAs based on the 14q32 locus. Using the signed averaged supervised method, two subgroups were defined within the MGH cohort with significantly different RFS stratified for the presence of metastasis at diagnosis (median RFS 100 months vs not reached and 13 vs 19 months, stratified log rank p=0.058), corroborating the previously hypothesized prognostic value of this miRNA signature (Fig. S2).

### Composite prognostic model integrating miRNA profiles and chemotherapy induced pathologic necrosis

Given that chemotherapy induced pathologic necrosis is the only currently utilized prognostic factor that has been proposed to therapeutically stratify patients, we reasoned that a composite model including both miRNA biomarkers and pathologic necrosis may provide increased and refined discriminatory power, as was suggested in the prior pilot study [7]. Indeed, when we created a composite model integrating both variables, we found that three separate patient subgroups were identified, with significantly different RFS (Fig. 3). Specifically, we identified a “very favorable”, a “very unfavorable”, and an “intermediate” outcome groups, suggesting that the molecular markers can be used synergistically with pathologic assessment to allow improved patient stratification. The “intermediate group” consisted of patients with good prognosis profile but suboptimal necrosis and vice versa, (these two subgroups were shown to not have different outcome, further justifying grouping them into one category). Confounding by metastatic status was also excluded as the distribution of metastatic or not metastatic tumors was not different between the three prognostic groups (Fisher’s p=NS). Similar clustering-based analysis for OS produced significant or trending results (Fig. S3).

**Figure 3.**
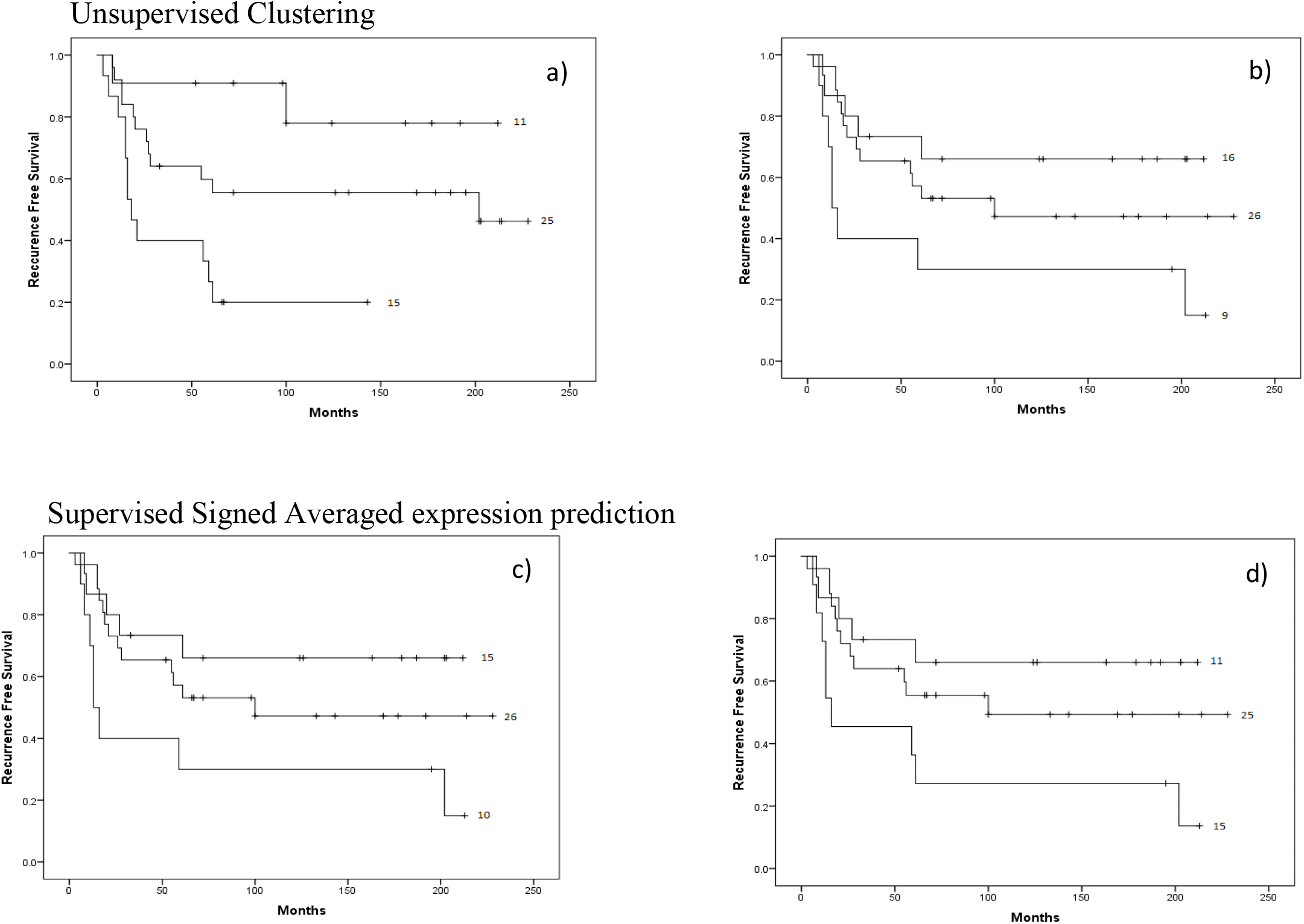
Kaplan-Meier log rank Recurrence Free Survival survival analysis of groups generated by a <u>composite classification</u> rule combining <u>miRNA profiles and pathologically assessed chemoresponse (PCR)</u>. Three groups were defined: “Very favorable” (good prognostic profile/optimal chemoresponse, top curve), “very unfavorable” (poor prognostic profile/suboptimal chemoresponse, bottom curve) and “intermediate” (good prognostic profile/suboptimal chemoresponse, or poor prognostic profile/optimal chemoresponse, middle curve). Clustering: a) 5 miRNA-profile/PCR, Median RFS 18 vs 202 months vs not reached, p=0.003. b) 22 miRNA-profile/PCR, Median RFS 21 vs 100 months vs not reached, p=0.026. Signed average: c) 5 miRNA-profile/PCR, Median RFS 13 vs 100 months, vs not reached, p=0.039. d) 22 miRNA-profile/PCR, Median RFS 16 vs 100 months vs not reached, p=0.039.

### Application of the prognostic miRNA profiles in the NCI TARGET osteosarcoma cohort

We used TaqMan miRNA RTPCR data provided through the recent NCI TARGET osteosarcoma cohort public release (n=86). After mapping and applying standard normalization, we performed unsupervised and supervised RFS analysis, using the same approaches as for the MGH sequencing cohort. Due to mapping attrition between the earlier generation microarray and TaqMan RTPCR platforms it was difficult to reconstruct the precise profiles, and the 5 and 22 miRNA profiles did not reach nominal significance. However, the pre-defined 27 miRNA profile including all the univariately prognostic miRNAs on 14q32, which mapped much more efficiently (26 of 27 probes) on the TaqMan platform, very closely approached prognostic significance. Applying a continuous risk score via the signed average supervised method we obtained a Cox regression, HR 1.32 (95% CI 0.98-1.78, p=0.065). The profile was also independent of confounding by metastatic status by multivariable Cox regression (HR 1.32, 95% 0.98-1.77, p=0.063). Furthermore, when controlling for metastasis at the time of diagnosis, clustering using the 22-miRNA profile (reduced to 18 miRNAs due to mapping attrition) showed a trend toward significance (median RFS 8 vs 27 months, and 64 vs 105 months for metastatic and non-metastatic subgroups, respectively, stratified log rank p=0.11). Of note, the NCI TARGET cohort has a much shorter follow up than both the MGH cohort and the previously published discovery cohort, potentially limiting its statistical power.

### Gene sets regulated by the miRNA profiles are strongly prognostic of recurrence and survival

To gain insight into the downstream consequences of the prognostic miRNA driven regulation, we analyzed the NCI TARGET mRNAseq data (large RNA sequencing, n=93) using experimentally verified gene targets for each of the prognostic miRNAs, identified through miRTarBase [13], a widely utilized database of miRNA target interactions. For each miRNA we compiled two gene target lists, an expansive list of experimentally verified targets and a restrictive list with additional targeted experimental evidence of miRNA/mRNA functional interaction.

We first performed Gene Set Analysis with the LS/KS test [14] for each list collectively for the end points of Recurrence Free and Overall Survival and found that several of the 5 and the 22 miRNA target gene sets were significantly prognostic of outcome (Table 2). The restrictive target list was too small (< 5 targets) for many of the miRNAs but still produced some significant results for miRNAs with enough gene targets included in the analysis. To further test for the possibility of random association due to the multitude of variables, we generated 10 random gene sets from the RNAseq data of equal size to the union of the 5 miRNA target lists (44 genes) and performed Gene Set Analysis. None of the random gene sets achieved the p value significance of the candidate miRNA target list by the LS test (Supplementary Methods and Results).

**Table 2.** miRNA target gene set analysis for RFS and OS with experimentally verified miRNA gene targets sets for the 22-miRNA profile (miRNAs from the 5-miRNA profile are shown in bold). All miRNA gene target sets with significant or trending LS permutation test p-values for the expansive analysis are shown.

| miRNA Gene Target Sets | Experimentally Verified Gene Target Lists |  |  |  |  |  |
| --- | --- | --- | --- | --- | --- | --- |
|  | Expansive | p-value (RFS) | p-value (OS) | Restrictive | p-value (RFS) | p-value (OS) |
| **hsa-miR-329-3p** | 344 | 0.00232 | 0.00552 | 8 | 0.00098 | 0.01705 |
| **hsa-miR-410-3p** | 157 | 0.00462 | 0.05839 | 10 | 0.09067 | 0.05576 |
| hsa-miR-139-5p | 69 | 0.00541 | 0.03526 | 25 | 0.01062 | 0.29243 |
| hsa-miR-335-3p | 193 | 0.02183 | 0.33236 | 3 | 0.34046 | 0.732 |
| **hsa-miR-487b-3p** | 15 | 0.07972 | 0.02288 | 8 | 0.07061 | 0.0044 |
| hsa-miR-432-5p | 63 | 0.08432 | 0.03707 | 4 | 0.54804 | 0.35184 |
| hsa-miR-493-5p | 70 | 0.09954 | 0.09239 | 3 | 0.34046 | 0.47078 |

Then, we performed unsupervised clustering and supervised signed averaged analysis with the union of the restrictive list gene targets from both the 5-miRNA (cluster R-index = 0.67) and the 22-miRNA (cluster R-index = 0.71) signatures on the NCI TARGET dataset and found that the gene targets consistently discriminated between two groups of samples with significantly different RFS (Fig. 4) and a strong trend for different OS (Fig. S4). Only one of the random gene lists demonstrated a prognostic discrimination as strong as that of the miRNA target gene list using the same methodology, suggesting that this finding was not a result of a random distinction due to a large number of possibly uninformative genes (File S1). Finally, we performed composite prognostic analysis with the gene target classification and pathologic necrosis, and as was the case for miRNA profiles, a similar distinction between three prognostic subgroups was observed (Fig. S5).

**Figure 4.**
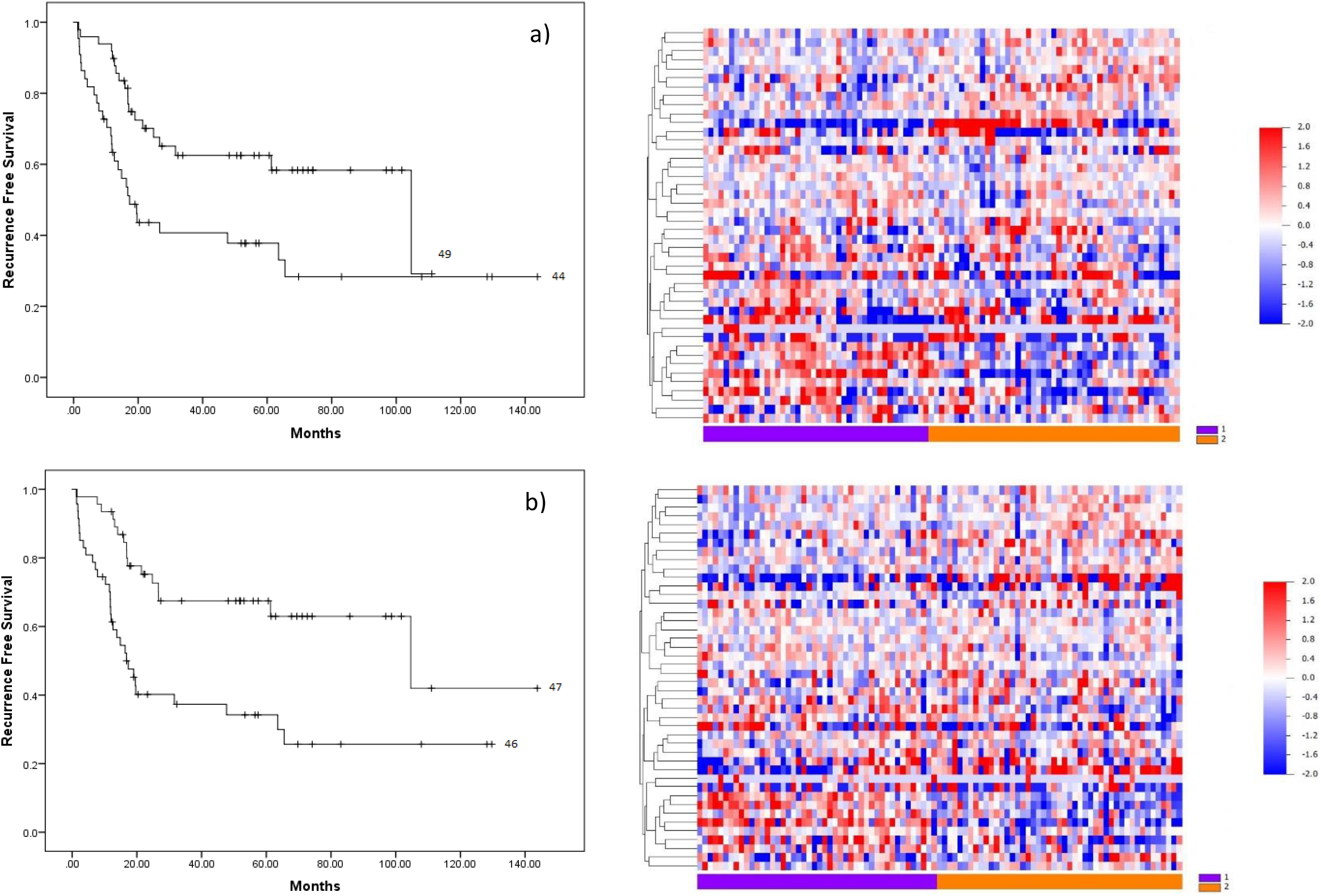
Kaplan-Meier Recurrence Free Survival analysis and gene target expression heatmaps of survival subgroups generated with the union of the restrictive list <u>gene targets of the 5-miRNA profile</u> for RFS. a) Unsupervised clustering, median RFS 17 (Group1) vs 105 months (Group2), log rank p=0.007. b) Supervised signed average prediction, median RFS 17 (Group1) vs 105 (Group2) months, log rank p=0.001.

### DNA methylation patterns of the miRNA prognostic signatures are also prognostic of outcome

We previously reported evidence for epigenetic regulation of the prognostic miRNAs and that their DNA methylation patterns correlate with miRNA expression and aggressive osteosarcoma behavior *in vitro* [8]. We tested the clinical relevance of this hypothesis using methylation array data from the NCI TARGET clinical cohort. We identified a total of 44 probes corresponding to CpG sites annotated to the 5-miRNA profile. We then assessed the methylation patterns of these probes in the two prognostic patient subgroups previously defined by 5-miRNA clustering and found that 15 of these CpG sites were differentially methylated between the two groups (p< 0.05). Further, the methylation pattern suggested that, on average, these CpG sites were hypermethylated in one group relative to the other (Table S2, Fig. 5a). Additionally, hierarchical clustering using the 44 methylation probes discriminated between two patient groups (R-index = 0.78) with strikingly different outcomes (Fig. 5b). To assess the robustness of this observation, we repeated the analysis after removing the 50% least variant CpG probes and observed that the survival results did not materially change.

**Figure 5.**
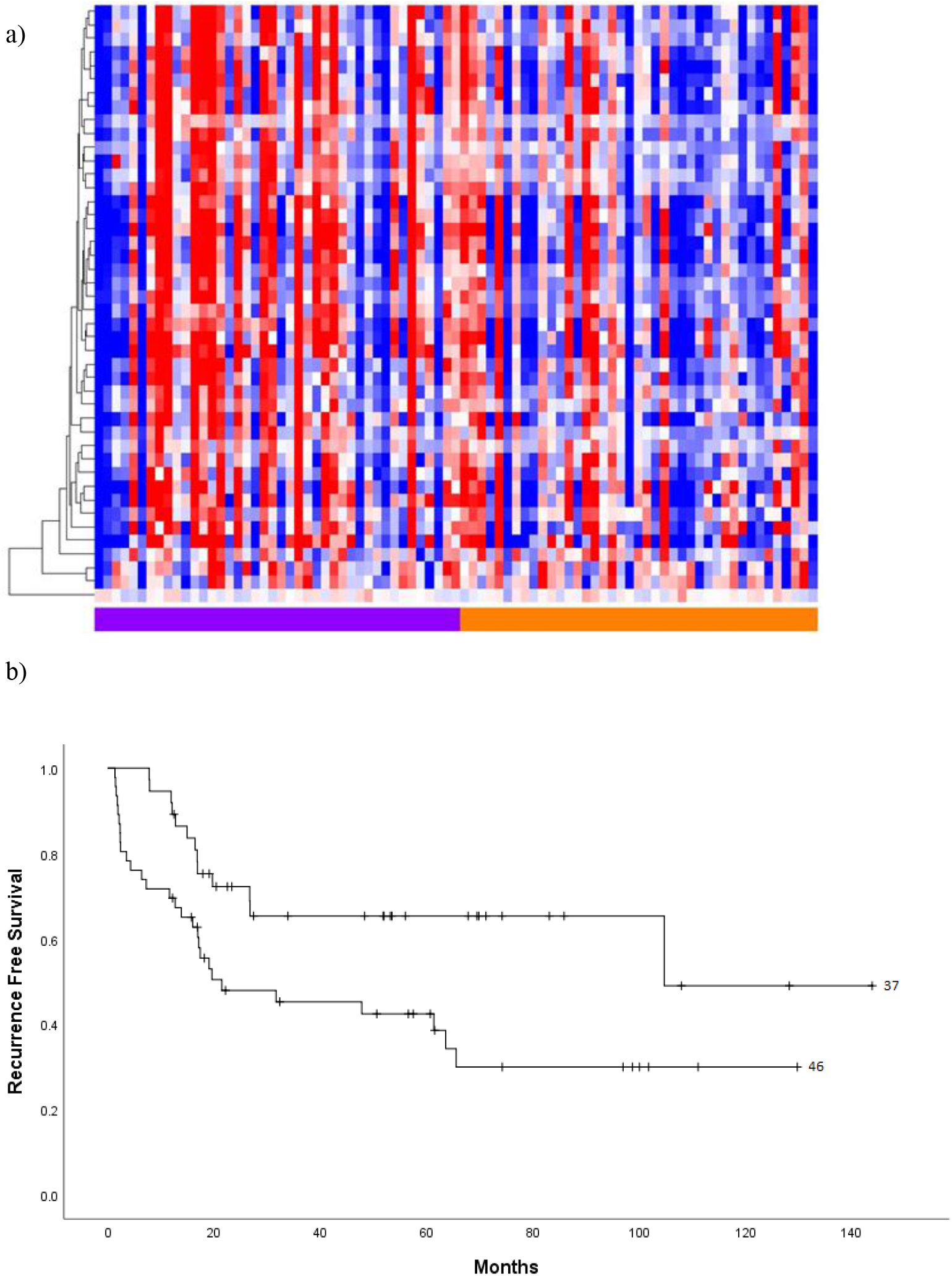
a) Heatmap depicting methylation differences between two groups defined by unsupervised hierarchal clustering using the 5-miRNA profile. b) Kaplan-Meier Recurrence Free Survival analysis of sample groups generated by unsupervised hierarchal clustering of the methylation probes annotated to the 5-miRNA prognostic profile. Median RFS 21 vs 105 months, log rank p=0.011.

Finally, we tested the association between DNA methylation and miRNA expression. We focused on the 4 of the 5 prognostic miRNAs located in the previously described, possibly epigenetically regulated non-coding cluster on 14q32. Each miRNA significantly correlated with at least 32 of the 44 CpG probes located in this genomic region (Spearman p <0.05 median Spearman coefficient 0.351, range 0.295-0.361, Table S3) suggesting miRNA patterns are partly modulated by DNA methylation.

### RNA networks define possible novel outcome related molecular phenotypes

We asked whether the transcriptomic patterns described above represent simple prognostic associations, or they may perhaps track underlying discernible phenotypes with possibly distinct molecular networks and clinical behavior. We reasoned that if there are wider molecular phenotypes hidden in the dataset, individual patient samples should be classified similarly as “high” or “low” risk using different types of molecular markers, mRNAs or miRNAs. We thus compared the miRNA-based and gene target-based classifications of the samples in the NCI TARGET dataset and found that they were significantly associated and highly concordant. For example, the Cramer’s V test for sample classification concordance between the 5 miRNA derived clusters and the 5-miRNA gene target derived (mRNA) clusters was 0.52 with a Fisher’s p<0.001 (detailed concordance findings in Table S4), irrespective of whether unsupervised or supervised methodology was used for classification. The survival associated subgroups generated using the 44 methylation probes annotated to the prognostic miRNAs were not concordant with the RNA based subgroups. These observations suggest that miRNA/gene target networks may track distinct molecular phenotypes of outcome in osteosarcoma, and that these phenotypes may be modulated by DNA methylation, which offers additional, but not fully overlapping prognostic information.

### Pathway analysis of miRNA targets

Building on the finding of the relationship between miRNA gene targets and outcome, we sought to identify associated molecular pathways with potential biologic or therapeutic implications. Gene set enrichment analysis was performed through the DAVID [15] functional annotation tool using the gene targets of the prognostic miRNAs. In order to strengthen the confidence in the detected pathways only BioCarta and KEGG [16] pathways with FDR < 0.05 [17] and EASE score p < 0.05 [18] in the restrictive 22 and 5-miRNA profiles, respectively, were considered significantly enriched. Using these criteria 29 KEGG and 10 BioCarta pathways were identified as enriched in both profiles (Table S5). Both the “PI3K” and “MAPK” pathways were identified as top-ranking enriched pathways, and 34 of the other pathways identified intersected “PI3K” or “MAPK” through key signaling enzymes such as PI3K, AKT/PKB, PTEN, MAPK1/3 or MEK. In order to test if these striking connections were specific to the identified pathways, we analyzed 10 BioCarta and 29 KEGG pathways with the lowest non-significant enrichment ranking within the 22-miRNA gene target set and only nine of them intersected the “PI3K” or “MAPK” pathways. Additionally, the “FOXO pathway”, downstream of AKT [19] and implicated in osteosarcoma [20], was identified in the BioCarta pathway analysis, and binding sites for FOXO1 were enriched in the genes of the miRNA targets through a parallel transcription factor binding analysis (FDR < 0.05). These results provide a glimpse into the downstream pathways which may become therapeutic targets in tumors with dysregulated miRNA expression.

### Pharmacogenomic integrative analysis reveals clinically applicable pharmaceuticals for osteosarcoma

The prognostic significance of the downstream miRNA gene targets prompted us to consider possible therapeutic applications of these findings. While screening for individual genes that could be targeted for specific inhibition can be useful, these large prognostic genomic signatures are often difficult to interpret biologically, and prioritization of a single gene or pathway is not always obvious. We opted for a comprehensive pharmacogenomic approach that analytically connects signatures, drugs and cell types aiming to prioritize novel therapeutic options for further development. We leveraged our PharmacoGx platform [21], and its web interface (PharmacoDB) [22], which provides for statistical analysis of primary experimental data from large publicly available databases of cell line drug sensitivities and gene signatures. We used PharmacoDB version 1.1.1 which includes 650,894 individual drug sensitivity experiments across 1,691 cell lines, and 19,933 possible gene markers currently included in the analytical pipeline.

In this analysis, we considered the top 20 experimentally verified gene targets (when available) for each of the 22 prognostic miRNAs ranked by their univariate p value for association with RFS. We input these genes into PharmacoDB, generating a list of 689 unique drugs for which any of the 415 gene targets were found to be a predictive marker of activity (one miRNA had only 15 available gene targets). Using our gene targets, PharmacoDB provided 340,436 experimental gene-drug correlations for further analysis. The drug list was filtered for reasonably large effect size (regression coefficient >|0.25|) and a stringent nominal p value (<0.001), with respect to association with at least one predictive gene marker, resulting in a list of 161 drugs. To increase the likelihood the observed drug effect relates to the miRNA network, we only considered drugs that produced at least three “hits” within the 22 miRNA/gene network and at least one “hit” within the 5 miRNA/gene network, resulting in a 57-drug list and further limited the list to 39 drugs with at least one interaction with 5 miRNA profile gene targets.

Then, we explicitly tailored the screening process to osteosarcoma. We obtained the median IC50 for the 34 out of the 39 drugs, for which PharmacoDB Batch Query provided drug response data for at least one of fifteen available osteosarcoma cell lines. We then set the IC50 for cisplatin, a core drug of active osteosarcoma regimens in clinical practice, as the minimum threshold for final drug selection, resulting in a final distilled list of 19 drugs (Table 3). Examining the final candidate list, we noticed that doxorubicin, another standard and probably the most active single conventional chemotherapy drug for osteosarcoma was included. Methotrexate and etoposide fell just below our stringent selection threshold but also produced “hits” when using slightly relaxed but still reasonably robust filtering criteria (coefficient>|0.15|, p < 0.05). These findings serve as a “positive control” that this analysis pipeline can discover active drugs, in addition suggesting that the miRNA networks may offer some predictive value for response to conventional chemotherapy. Further, sixteen of the 19 drugs in the final list have been included in phase I or phase II clinical trials for the treatment of various types of cancer.

**Table 3.** Pharmaceuticals for which PharmacoDB analysis reveals a drug response predictive association with miRNA gene targets. Methotrexate, etoposide, and cisplatin are included for comparative purposes. Median IC50 values are obtained across all osteosarcoma cell lines. The median regression coefficient is calculated from the significant gene – drug interactions used to identify the drugs. The nine bolded drugs act through pathways enriched in target genes of the miRNA profile (shown and bolded in Table S5). Specifically, dinaciclib, CGP060474, and alvocidib inhibit the “cell cycle pathway”, trametinib, AZ628, and PD-0325901 inhibit the “MAPK pathway”, dasatinib and nilotinib act through the “Inhibition of Cellular Proliferation” by Gleevec pathway, and pictilisib inhibits the “PI3K pathway”.

| Name | Drug Class | Median IC50 (μM) | Median Regression Coefficient | Chemical Replicates |
| --- | --- | --- | --- | --- |
| thapsigargin | SERCA inhibitor | 0.003 | 0.273 | 1 |
| daporinad | NAPRT inhibitor | 0.004 | 0.323 | 1 |
| paclitaxel | microtubule stabilizing agent | 0.016 | 0.552 | 1 |
| **dinaciclib<sup>†</sup>** | **CDK inhibitor** | 0.016 | 0.288 | 3 |
| **CGP-60474<sup>†</sup>** | **CDK inhibitor** | 0.028 | 0.266 | 3 |
| panobinostat | HDAC inhibitor | 0.112 | 0.291 | 1 |
| WH-4-023<sup>†</sup> | Src inhibitor | 0.156 | 0.268 | 2 |
| methotrexate | DHFR inhibitor | 0.160 | - | 1* |
| **alvocidib<sup>†</sup>** | **CDK inhibitor** | 0.221 | 0.283 | 3 |
| irinotecan | topoisomerase inhibitor | 0.444 | 0.265 | 4* |
| doxorubicin | topoisomerase inhibitor | 0.521 | 0.260 | 4* |
| topotecan | topoisomerase inhibitor | 0.698 | 0.273 | 4* |
| **trametinib** | **MEK inhibitor** | 0.897 | 0.270 | 2 |
| **AZ628<sup>†</sup>** | **RAF inhibitor** | 0.966 | 0.256 | 1 |
| TAE684 | ALK inhibitor | 1.143 | 0.266 | 1 |
| **dasatinib** | **Abl/Src inhibitor** | 2.846 | 0.304 | 2 |
| **pictilisib** | **PI3K inhibitor** | 3.359 | 0.452 | 1 |
| etoposide | topoisomerase inhibitor | 3.678 | - | 4* |
| tozasertib | aurora kinase inhibitor | 4.504 | 0.377 | 1 |
| **PD-0325901** | **MEK inhibitor** | 5.199 | 0.292 | 2 |
| **nilotinib** | **Abl inhibitor** | 5.703 | 0.278 | 1 |
| cisplatin | alkylating agent | 8.414 | - | 1* |

\* The drug class includes a chemical replicate corresponding to a reference pharmaceutical.
<sup>†</sup> Experimental drug response (IC50) data is only available for a limited number ( $\leq 3$ ) of osteosarcoma cell lines

Of these, four drugs are part of standard care, four are targeted agents that have or are being tested in dedicated sarcoma trials, including panobinostat [23], alvocidib [24], dasatinib [25], and nilotinib [26],[27], and two of which have been tested in osteosarcoma; namely alvocidib and dasatinib, which has shown some evidence of activity in an early clinical trial. In this trial, the 46 high-grade, metastatic, pre-treated, osteosarcoma subjects experienced a 15% two-year survival rate, 11% experienced 6-month progression free survival, and three subjects experienced >10% decrease in tumor size, or >15% decrease in tumor density [25]. Despite the modest result observed with dasatinib monotherapy in the SARC009 trial, our analysis predicts for potential synergy between dasatinib and MAP chemotherapy in a subset of patients undergoing standard treatment. To this end, dasatinib has previously been shown to synergize with doxorubicin *in vitro* [28]. Finally, a recently published clinical trial of regorafenib vs placebo, (SARC024 [29]), reported increased progression free survival for pre-treated, metastatic osteosarcoma patients. We thus tested if there is an interaction between the miRNA target profile and regorafenib response and we discovered that this indeed is the case, using the previously described cut offs (coefficient >|0.15|, p<0.05). Of note, further support for *in vitro* synergy between regorafenib and doxorubicin, has been provided in a different cancer type [30].

Six other non-classic chemotherapeutic drugs from the list have been tested in clinical trials, though not yet with osteosarcoma patients. For these drugs we also found *in vitro* data for effect in osteosarcoma cell lines, separate and independent of the experiments already included in PharmacoDB. These drugs include dinaciclib [31], panobinostat [32], trametinib [33], TAE684 [34] (ceritinib), pictilisib [35], and tozasertib [36]. Interestingly, TAE684 (ceritinib) was previously found to reverse cisplatin resistance *in vitro* in osteosarcoma by our own sarcoma research group [34]. Recently, clinical efficacy for nilotinib in high-risk chordoma subjects (another type of primary bone tumor) was established by our own sarcoma research group, where the median overall survival time was 61.5 months [26]. Additionally, a phase II trial for single agent panobinostat was prospectively registered in March of 2018, which is designed to include a sub-cohort of 20 osteosarcoma patients (ACCT008/NORTH). A phase I/IIa trial studying trametinib is recruiting patients, including with osteosarcoma (NCT02124772). These proof-of-principle findings suggest this approach can identify compounds with at least initial evidence for activity against osteosarcoma and, additionally, could serve as a large-scale screening tool to prioritize use of these compounds based on tumor profiles of candidate miRNA/mRNA networks presented in this study.

We finally examined the intersection of the results of the pharmacogenomic analysis and the analysis of the pathways enriched in the miRNA target sets (Tables 3 and 4). We found that four of the drugs identified in this pharmacogenomic analysis directly target the MAPK or PI3K pathway, both of which are enriched in the gene target set of the prognostic miRNAs. Because aberrant miRNA expression appears to modulate these pathways, inhibiting key elements may be especially effective in tumors where the miRNA signatures predict poor prognosis. Therapeutics targeting these pathways have shown *in vitro* activity in osteosarcoma, and ongoing clinical trials (NCT03458728 and NCT02124772) are evaluating drugs targeting both mechanisms, one of which includes combination therapy with trametinib, a drug identified in our analysis.

## DISCUSSION

Osteosarcoma treatment is based on a combined modality approach consisting of surgery and neoadjuvant and adjuvant chemotherapy with the MAP regimen. This treatment standard, unchanged in the last 20-30 years, has led to an improvement in outcome compared to poor results previously obtained with surgical resection alone, but leaves much to be improved, as up to 50% of the patients, many of them children and young adults, eventually relapse and succumb to their disease. Development of new therapies is challenging because of the relative rarity of the tumor and the lack of robust biomarkers that would help stratify outcome risk and prioritize patients for trials. In this context, we, and others, have previously reported initial findings suggesting that miRNA profiles, obtained at the time of pre-chemotherapy diagnostic biopsy, can be promising biomarkers of outcome following standard therapy[7],[8],[37]. These initial reports were based on relatively small sample sizes and expression microarray technologies with inherent technical limitations and uncertainty about their wider reproducibility. In this study, we tested hypotheses based on miRNA profiles and their associated gene target networks, validated their prognostic power in two large genomic datasets, and placed them in the wider molecular context of relevant tumor outcome phenotypes in osteosarcoma.

Our two study cohorts (n=74, 95) are large for this rare disease and such sample sizes have not been reported to date in osteosarcoma biomarker studies. Our study is unique in that it combines the strength of a large well annotated cohort based on a single expert referral center, (Massachusetts General Hospital) with presumably more limited clinical heterogeneity, with the strength of a large multicenter, nation-wide cohort, based on the NCI TARGET initiative, capturing real life heterogeneity and variability of a large population across the entire North American continent. Two different genomic assays (small RNA sequencing and TaqMan miRNA qRTPCR) were used in the two cohorts, and messenger (large) RNAseq and methylation array data were also available for one cohort. Despite these inherent challenges in the analysis and validation process across multiple platforms, our study findings support the hypothesis that these miRNA profiles and networks can be reproducible across a large external population, bringing them one step closer to eventual clinical applicability.

We used simple methodology (signed averaged expression) to calculate risk scores, an approach that is inherently less prone to overfitting, as it does not rely on identifying an optimal model. In addition, unsupervised clustering analysis, which is practically devoid of overfitting also resulted in significant survival discrimination, further supporting the robust prognostic information carried by the miRNA profiles. That said, when a single technology is eventually chosen for clinical application, it is conceivable that a fully parametric multivariate model may be optimal for individualized outcome prediction.

Particularly notable is the fact that sets of experimentally validated gene (mRNA) targets of the prognostic miRNAs were also found to be strongly prognostic of outcome suggesting a possibly active miRNA/mRNA network. Furthermore, we observed a significant and moderately strong concordance between miRNA and mRNA classification of individual samples as ‘high” or “low” risk. This despite the inherent variability of these analyses and the fact that different assays were used for miRNA and mRNA analysis in the NCI dataset. This observation supports the notion, initially suggested in our early studies, that miRNA patterns are not simply markers of outcome but may track underlying molecular phenotypes where distinct miRNA/mRNA networks modulate tumor behavior and patient course.

To the extent that relevant data were available, we also demonstrated that the prognostic value of the miRNA profiles was independent of the main known clinicopathologic variables that affect outcome in osteosarcoma, namely the presence of metastatic disease at the time of diagnosis and pathologic necrosis in response to neoadjuvant MAP based chemotherapy. Furthermore, we propose a simple composite model integrating information from both pathologic necrosis and miRNA biomarkers that allows improved and more refined stratification into three relevant prognostic groups, again validating a hypothesis previously suggested in our pilot work [7]. Given the failure thus far to improve patient outcomes based on conventional “high” and “low” risk stratification using pathologic necrosis, a three-group composite stratification approach may offer a more powerful and refined application. Based on this model, a very favorable prognosis group scan be safely monitored with a negligible risk of recurrence. An intermediate prognosis group can be approached by selectively developing new treatments adding to the backbone of standard MAP chemotherapy. Finally, a very unfavorable prognosis group may derive a very limited benefit from standard chemotherapy. In our analysis, this group appears to have an outcome equal to what was historically the case for surgical treatment in the pre-chemotherapy era and may require an entirely different approach to test novel therapies based on the unique molecular subtype that this group represents.

The finding that DNA methylation patterns corresponding to the prognostic miRNAs are also prognostic of outcome provides large scale clinical validation of tentative observations made in prior pilot small sample cohorts [8],[38]. In addition, the correlation between methylation and miRNA expression implies that there is a potential for a regulatory mechanism, at least in the 14q32 non-coding locus, which hosts most of the prognostic miRNAs. However, the sample classification between the methylation and RNA markers was discordant suggesting that the methylation patterns are not simply duplicating information contained in the transcriptome. It is not clear at this point, whether the specific methylation markers reported here are uniquely relevant to miRNA expression and outcome, or whether they are surrogates for more global methylation patterns in osteosarcoma, as was recently suggested [38].

Through our work and that of others, the prognostic value of miRNA profiles in osteosarcoma appears to be validated. Clinical application of these profiles will further rest in their potential to guide treatment. The recent large randomized international EURAMOS trial, failed to detect a survival advantage to inclusion of ifosfamide/etoposide to the post-operative regimen for patients with poor pathologic response [6], as well as inclusion of interferon a for the patients with good pathologic response [3]. These findings may signify the lack of true benefit from the utilized drugs, but alternatively may also be partly due to the possibility that pathologic necrosis after 10 weeks of MAP treatment may not be the optimal marker, in isolation, for stratifying patients for alternate treatment. Our composite prognostic analysis combining pathologic necrosis and miRNA profiles offers a possible new powerful and refined method to stratify patients for various treatment strategies.

Our pharmacogenomic approach coupled with pathway analysis provides initial insights into novel therapeutic strategies, whereby new drugs, that may be already developed for other indications, or are shown to have activity against osteosarcoma in clinical trials (such as dasatinib [25], regorafenib [29]) can be shown to be active or synergistic in patients who are receiving standard MAP chemotherapy based on the miRNA network profiles. While these new paradigms require additional validation in both clinical as well as experimental studies, this approach to drug repurposing is particularly well suited for this rare tumor where drug development has stalled for the last 30 years. Several possible drug candidates are revealed, including CDK inhibitors, MEK inhibitors, as well as PI3K pathway inhibitors, the therapeutic value of which was also proposed by a recent osteosarcoma genomic study in a different clinical and experimental setting [39]. Our data further support these findings and suggest that miRNAs may partly modulate the PI3K effect in a subset of osteosarcoma tumors and may allow stratification for the use of relevant inhibitors in clinical practice.

While the findings on the MGH dataset more closely resembled and validated hypotheses initially presented in the early studies, the findings from the TARGET miRNA expression data were suggestive of a prognostic association but did not reach the same level of definitive significance. This may be due to several confounding issues such as the less complete clinical annotation and shorter follow up of the TARGET dataset, differences between the cohorts, such as in median age (22.5 vs 15 years), and differences in assay technologies used (RNA seq vs RTPCR) and related cross platform probe mapping attrition. Nonetheless, the strong prognostic value of the miRNA gene targets in the NCI TARGET data (derived via RNA seq technology from larger patient cohort than the miRNA assays) and the fact that the profiles in the NCI TARGET data show a strong trend for significance with stratification for metastatic disease, strongly supports the notion that miRNA derived networks do have prognostic influence and this may become more clear when longer follow up and more complete annotation, particularly full treatment information, becomes available from the NCI.

Future functional studies will help elucidate the precise mechanisms by which the miRNA/mRNA networks affect tumor behavior and how best to exploit them therapeutically in a specific manner, as initial reports have suggested [40]. Further clinical steps may include testing the possibility that the molecular markers or phenotypes statistically interact or predict for an effect from the use of intensified or alternative chemotherapy, such as ifosfamide/etoposide, which remains an active regimen despite the failure to demonstrate survival benefit in the EURAMOS [6] trial. In addition, the use of these markers via pharmacogenomic analysis as a tool to select patients for clinical trials of new candidate drugs merits further testing. Finally, from a research strategy perspective, our study exemplifies the benefit of maximizing use of local resources in a large single institution referral center coupled with NCI supported efforts to generate national cooperative group-based resources and molecular data that would be impossible to otherwise efficiently generate and disseminate. We believe this is a great model to accelerate scientific discovery and clinical validation, particularly in rare or pediatric tumors.

## CONCLUSIONS

We performed the first small RNA sequencing osteosarcoma study to date and validate candidate prognostic miRNA signatures associated with outcome independent of known clinicopathologic factors. Furthermore, integration of a large, multi-omic, national cooperative group supported dataset revealed that downstream regulatory targets of the miRNA signatures are also prognostic of outcome, suggesting that miRNA driven molecular networks represent previously undefined outcome phenotypes in osteosarcoma. These subtypes may be translatable for stratifying patients for novel and alternate therapeutic strategies.

## METHODS

### The MGH human osteosarcoma cohort RNA isolation and small RNA sequencing

Eighty (80) pre-chemotherapy diagnostic frozen samples from patients with high grade osteosarcoma were retrieved from the MGH Pathology archives based on an IRB approved retrospective tissue and clinical information protocol. Samples were banked between 1994-2013 and selection was done chronologically, subject only to tissue availability, pathology confirmation, and confirmation of treatment with neoadjuvant and/or adjuvant chemotherapy. All slides were reviewed again by an expert study pathologist confirm diagnosis and to determine tumor cellularity. Six samples with very low tumor cellularity (<5%) were excluded resulting in 74 samples selected for further study.

RNA was isolated with the QIAGEN RNeasy® Plus Universal Mini Kit (Qiagen) using an adapted protocol designed to increase the quality of extracted RNA from bone [41]. Sequencing was performed at the Center for Cancer Computational Biology at Dana Farber Cancer Institute (Boston, MA). RNA quantity was determined on the Qubit using the Qubit RNA High Sensitivity Assay Kit (Life Tech) and RNA quality was determined on the Bioanalyzer using the RNA Pico Kit (Agilent). Using the NEBNext Multiplex Small RNA Library Prep Kit for Illumina (NEB), 100ng of total RNA was converted into a DNA library following the manufacturer’s protocol, with no modifications. Library quantity was determined using the Qubit High Sensitivity DNA Kit (Life Tech) and library size was determined using the Bioanalyzer High Sensitivity Chip Kit (Agilent). Finally, libraries were subjected to qPCR using the Universal Library Quantification Kit for Illumina (Kapa Biosystems) and run on the 7900HT Fast qPCR machine (ABI). Libraries passing quality control were diluted to 2nM, combined into library pools, and sequenced on the NextSeq 500 (Illumina) at a final concentration of 2pM. Sequencing was done across three NextSeq Single Read 75 Cycle High Throughput V2 flowcells following standard protocols. Small-RNAseq reads were processed and quantified using the sRNAtoolbox sRNAbench tool [42]. The pipeline automatically identified and removed adapter sequences from the input fastq files.

Hierarchal read mapping was performed to first map reads to the UniVec database to filter common laboratory contaminants, and then to rRNAs. The preprocessing steps are performed with Bowtie [43]. The remaining reads are then mapped to human miRNAs annotated in miRbase [44] version 21. Mature and precursor miRNAs were independently mapped and quantified. The raw mature miRNA read counts (2003 transcripts) were normalized and log base 2 transformed using the DESeq2 R package [45] prior to analysis.

### NCI TARGET human osteosarcoma cohort and associated molecular data

A publicly available human osteosarcoma molecular dataset was obtained from the NCI TARGET (Therapeutically Applicable Research to Generate Effective Treatments) osteosarcoma project data matrix (https://ocg.cancer.gov/programs/target/data-matrix). This dataset currently (data collection is ongoing by the NCI) contains a total of 95 clinically annotated samples for which ABI TaqMan Megaplex^™^ human miRNA qRTPCR data (ABI) (n = 86), Illumina RNA seq data (n = 93), and Illumina Infinium HumanMethylation450K (n = 83) is available. Experimental and data processing methodology for miRNA expression, RNA seq, and methylation data, employed by the TARGET study team, is available on the TARGET osteosarcoma webpage (https://ocg.cancer.gov/programs/target/target-methods). We transformed and normalized raw miRNA qRTPCR data using the standard 2^-ΔCt^ transformation as previously described [46]. We further analyzed the gene level RNAseq data provided by TARGET. All transcript reads less than 200 bases in length based on the Ensembl Genome Reference Consortium Human Build 38 patch release 12 (GRCh38.p12) were removed. DESeq2 [45] was then used to normalize and log base 2 transform the read count expression data. Gene annotations were obtained through the Ensembl BioMart tool [47]. Beta mixture-quantile normalized [48] methylation β values were downloaded, and then inter-array normalization and M value [49] conversion was performed via the *methylumi* R package [50]. HumanMethylation450 v1.2 manifest file was used to obtain probe annotations and is available at https://support.illumina.com/downloads/infinium_humanmethylation450_product_files.html.

### Candidate signature mapping across experimental assays and platforms

We mapped candidate prognostic miRNAs from the previously published discovery dataset, generated using DASL miRNA microarrays [7], to mature human miRNAs in the MGH RNA sequencing dataset and the NCI TARGET qRTPCR dataset, utilizing miRbase [44] and the BLASTN algorithm. We only considered for further analysis miRNAs with perfect 18-mer contiguous sequence homology to the original DASL probes (some of which were only 18 nucleotides long). This mapping selected 21 of the original 22 prognostic miRNAs on the MGH dataset, and 18 of the 22 miRNAs on the NCI TARGET dataset. In addition, for the 27 miRNA pre-defined prognostic profile on the 14q32 locus [8], mapping selected 27/27 miRNAs on the MGH RNA seq cohort and 26/27 miRNAs on the TARGET TaqMan RTPCR cohort. Detailed mapping output is shown Table S1.

### Unsupervised and Supervised Survival analysis and prediction

Unsupervised hierarchal clustering using the previously described prognostic miRNA profiles in the MGH and TARGET datasets was performed with the centered correlation and average linkage method [51] and resulting groups were then analyzed for survival differences. Cluster reproducibility was assessed with the R-index [12]. For supervised survival analysis, we used the simple and robust signed average method as previously described by us and others [8],[52],[53],[54]. In this approach, the expression of the miRNAs in a given prespecified profile was averaged and weighted by +1 or -1 depending on the direction of the Hazard Ratio/coefficient of each miRNA’s association with outcome on a univariate Cox Regression analysis. Signs were taken from the previously published pilot study. Using this metric as a prognostic risk index two groups were defined by the median value as a cut off. Survival differences between cluster defined or signed average defined groups were tested with Kaplan-Meier analysis and the log rank test for significance. Multivariate analysis for confounding prognostic factors was performed using a Cox Regression model, with the profile and relevant factors entered as independent variables.

### Bioinformatic identification miRNA gene targets

miRTarBase [13] was used to identify experimentally verified gene targets of the miRNA profiles. The miRTarBase reports miRNA gene targets at different levels of confidence depending on the type of experimental assays (reporter assay, Western blot, qPCR, microarray, NGS, pSILAC (pulsed Stable Isotope Labeling by Amino Acids in Cell Culture). We defined two gene target lists, one “restrictive” and one “expansive” based on a higher and a relatively lower confidence for miRNA-mRNA functional interaction, respectively.

### Differential expression, Gene set analysis, and Standard statistical tests

Continuous variable differential analysis between two groups of samples was performed by a t-test with p-values adjusted to control the false discovery rate (FDR) using the Benjamini – Hochberg step-up procedure for multiple testing [55]. Gene set analysis for the association of the expression profiles of miRNA gene targets with survival was performed with the functional class scoring method, applying the LS/KS test with appropriate permutation-based p values [14]. For the expansive miRNA gene target analysis, a filter was applied to remove genes with the lowest 20% variance, to reduce statistical noise. Associations between two categorical variables were evaluated with two-tailed chi-square/Fisher’s exact test. Cramer’s V test employed to assess the strength of the classification concordance between different profiles. Spearman rank and Pearson’s r statistics were used to evaluate continuous variable correlations.

### MiRNA gene target pathway enrichment analysis

Enrichment of BioCarta pathways, KEGG [16] pathways, and transcription factor binding sites in the prognostic miRNA gene targets was evaluated using the DAVID Functional Annotation Tool [15]. BioCarta and KEGG pathways with FDR [17] < 0.05 and EASE score [18] p < 0.05 in the restrictive 22 and 5-miRNA gene target lists, respectively, were considered significant. Many KEGG terms generic to cancer were identified as significant. We considered these terms to be redundant, and less specific than other pathways, and thus omitted them from the results, but the complete results are available in File S1. For transcription factor binding site analysis, there is still significant uncertainty about experimentally verified annotations, and a lot of nominally significant sequences were identified, so an equally stringent significance cut off of FDR < 0.05 was used in both 22-miRNA and 5-miRNA restrictive gene target lists.

### Integrative Pharmacogenomic Analysis

We used the miRNA gene target profiles for pharmacogenomic drug interaction discovery using PharmacoDB [22] interface, which performs data analysis via the PharmacoGx R package [21]. The expansive list of experimentally verified gene targets of the 22-miRNA profile was ranked by univariate p-value for association with RFS. The 20 top gene targets for each miRNA were entered in the PharmacoDB and PharmacoGx pipeline analyzing drug/gene predictive interactions across seven large scale datasets including a total of 650,894 individual drug sensitivity experiments across 1691 cell lines, and 19933 possible gene markers via a multivariate regression model adjusting for tissue source and experimental batch. We chose drug candidates at a stringent 0.001 regression p-value for predictive association with gene markers, and a regression coefficient > |0.25| for effect size. To increase the specificity of the resulting drug list, only drugs passing the p-value and effect size filters for at least three of the 22-miRNA profile gene targets and at least one of the 5-miRNA profile gene targets were retained for further analysis. The *in vitro* experimental data of the filtered drug list was then evaluated specifically in 15 osteosarcoma cell lines through the PharmacoDB Batch Query to obtain sensitivity measures (IC50 dose response metric) for response to candidate drugs. For each drug we calculated the median IC50 across all cell lines to minimize outlier effects and drugs were ranked by median IC50 value. The final drug sensitivity list was compared to the IC50 values for standard chemotherapeutic drugs used and known to be effective in osteosarcoma.

### Statistical Software

The NCI BRB-ArrayTools v4.6.0 [56], R (version 3.4.3), and SPSS v 24 software, were used.

## Data Availability

The independent NCI TARGET datasets are available through the data matrix at https://ocg.cancer.gov/programs/target/data-matrix .

## ADDITIONAL FILES

**Supplementary Methods and Results**. Additional details of the methodology and results are presented. (DOCX)

**Figure S1**. Kaplan-Meier log-rank Overall Survival analysis of candidate profiles on the MGH dataset by the supervised signed average prediction method. (PNG)

**Figure S2**. Kaplan-Meier log-rank Recurrence Free Survival analysis using the 27-miRNA profile and the signed averaged supervised method defined subgroups stratified for the presence of metastasis at diagnosis. (PNG)

**Figure S3**. Kaplan-Meier log rank Overall Survival analysis of groups generated by a composite classification rule combining miRNA profiles and pathologically assessed chemoresponse (PCR). (PNG)

**Figure S4**. Kaplan-Meier analysis of Overall Survival groups generated by unsupervised clustering and supervised signed average analysis with the union of the restrictive list gene targets of the 5 and 22 miRNA signatures on the NCI TARGET data. (PNG)

**Figure S5**. Kaplan-Meier log rank Recurrence Free Survival analysis of groups generated by a composite classification rule based on miRNA target gene profiles and pathologically assessed chemoresponse (PCR). (PNG)

**Table S1**. 5-miRNA and 22-miRNA profiles (and the secondary 27 miRNA profile) mapped to each dataset. (DOCX)

**Table S2**. Differential methylation of the CpG probes annotated to the 5-miRNA profile, between the miRNA defined cluster groups. (DOCX)

**Table S3**. Median Spearman coefficients comparing 5 miRNA profile (the 4 located on 14q32) expression and methylation. (DOCX)

**Table S4**. Cramer’s V and Fisher’s exact test results for concordance between the classifications derived from the miRNA signatures and the corresponding gene targets. (DOCX)

**Table S5**. All significantly enriched BioCarta pathways, KEGG pathways, and transcription factor binding sites in the prognostic miRNA gene targets. (DOCX)

## DECLARATIONS

### Ethics approval and consent to participate

Samples were selected based on an IRB approved retrospective tissue and clinical information protocol.

### Consent for publication

Not applicable.

### Authors’ contributions

DS, JQ, WTB, GPN, EC, DE, FJH, YLC, SLC designed the study.

CL, CG, YW, BL performed experiments and generated data.

CL, CG, DS, YW, BL, GC, ZD, WTB, VD, BHK, analyzed and interpreted data.

CL, DS, WTB, BHK, wrote the manuscript.

## Acknowledgements

We wish to thank Haotong Wang, Ruoyu Miao, and Jason Kim, for assistance with retrieving clinical data, Christie Swett for assistance with specimen retrieval, and Renee Rubio for technical assistance with sequencing. The results presented here are in part based upon data generated by the Therapeutically Applicable Research to Generate Effective Treatments (TARGET) initiative, which is publicly available at https://ocg.cancer.gov/programs/target/data-matrix. Information about TARGET can be found at http://ocg.cancer.gov/programs/target.

